# Pseudoscience in Cancer Services; a survey of National Health Service Trusts in England

**DOI:** 10.1101/2024.06.26.24309516

**Authors:** Leslie B Rose

## Abstract

**Background:** Scientifically implausible treatments are offered by some hospital cancer departments. Examples are reiki, aromatherapy, and reflexology. Salaried practitioners are employed to deliver these therapies, which are provided as palliative care, although they lack evidence of effectiveness. Such practices seem to conflict with efforts to make health care evidence based.

**Aim:** To estimate the extent of certain pseudoscientific practices in NHS Trusts, and to evaluate the rationale for such provision.

**Design:** Relevant documents were requested from NHS Trusts under the Freedom of Information Act 2000 (FOIA). Main outcome measures were: number of trusts offering pseudoscientific practices in cancer departments, time to full FOIA response, presence and content of practice governance documents, and presence and quality of evidence for practices.

**Setting/Participants:** Cancer care departments in NHS hospitals in England. No patient participants were involved in the survey.

**Results:** 13.6% of eligible NHS trusts were offering pseudoscientific clinical practices. No trust provided a valid business case, or any robust evidence for the practices. The governance documents included claims about chakras, meridians, and invisible “energy”. Ten trusts required that informed consent be obtained from patients. This could not have been obtained because information given was misleading.

**Conclusions:** Pseudoscientific practices are embedded in the NHS in England, and governance documents show poor understanding of clinical evidence.

## Introduction and Background

The NHS has made considerable strides in recent years to embed evidence based medicine in clinical practice [1]. Obviously ineffective practices such as homeopathy are no longer funded [2]. However, some specialisms not only retain other long-established practices for which no robust evidence exists, but regularly introduce so-called complementary therapies based on implausible ideas about how the body works. This distinguishes them from scientifically plausible but poorly evidenced modalities.

The term pseudoscience is applied to claims that are not based on science, but on imaginary mechanisms. For example, physiotherapy is an established discipline with a plausible scientific basis. However, approximately half of physiotherapists do not choose treatments with robust evidence of effectiveness [3]. In contrast, anthroposophical medicine was invented by Rudolf Steiner, and is based on mystical concepts including astrology, reincarnation, and sympathetic magic [4]. Physiotherapy is not pseudoscience, but anthroposophical medicine is.

In August 2022 articles appeared in the national press about recruitment advertisements for a reiki practitioner in an NHS palliative care unit [5, 6]. Reiki is a pseudoscientific practice in which practitioners claim to be able to sense and manipulate a patient’s “vital force” or invisible “energy”, with benefits claimed for health and well-being. It has been extensively researched, and no consistent and robust evidence to support the claims has emerged [7,8].

The national press articles identified that reiki practitioners were being funded by a charity, The Sam Buxton Sunflower Healing Trust. The charity’s website carries a list of NHS units which have been beneficiaries of this programme [9]. Other pseudoscientific practices have been encountered as being used in the NHS, of which not all were charity funded.

The aim of this study was to estimate the prevalence and evaluate the offer/provision of pseudoscientific therapies used in the NHS.

## Methods

### Search for NHS units offering pseudoscientific therapies

To find NHS units potentially offering reiki, all instances of websites with URLs ending in nhs.uk were found using the following search in Google: site:nhs.uk reiki

Websites describing cancer services, including oncology and palliative care, were selected from the search results. The survey was not intended to be confined to England, but the search did not provide positive results for other UK nations. The number of NHS trusts in England was identified as 2290 [10], of which 10 are ambulance trusts and 50 are mental health trusts (ie 169 relevant trusts for the survey). General practices are not public bodies so were not included in the survey.

Having started with NHS units offering reiki, the search was extended to other pseudoscience modalities. The search command was repeated after modifying and replacing “reiki” with each of the following terms: Aromatherapy, EFT (emotional freedom technique), Reflexology, Chakra balancing, Therapeutic touch, Indian head massage, Bach flower remedies.

### Requests for Information using FOIA

A pilot request was made to Manchester University NHS Foundation Trust on 15^th^ August 2022, in respect of their vacancy for a reiki practitioner.

Freedom of Information Act 2000 (FOIA) requests were then made to 12 further NHS units identified by the Google searches as providing reiki. Requests were made between 30^th^ August - 22^nd^ September 2022.

In total, 13 trusts (including Manchester), were asked to provide:

1. All correspondence with the Sam Buxton Sunflower Healing Trust
2. All internal correspondence relating to the position advertised
3. All internal procedures and guidance governing the creation of the position advertised.

A third round of requests was made to nine more NHS units who provided any of the therapies identified by the expanded search. The requests were made from 17^th^ September to 2^nd^ October 2022, asking them to provide:

1. The business case for providing the therapy, and the clinical evidence on which it is based.
2. Internal correspondence and meeting minutes which document the decision to provide the therapy.
3. The internal guidance governing the provision of the therapy.
4. The justification for the mode of action claimed.

Two requests were sent each to University Hospitals Bristol and Weston NHS Foundation Trust, and to University Hospitals Dorset NHS Foundation Trust; the first about reiki, and the second about reiki and other practices. This was because other practices were identified later.

In order to improve the response rate, late responders (defined as a trust not responding within 20 working days) were followed up firstly by making a request for an internal review of the public body’s failure to respond (as per FOIA policy), and then, by a complaint to the Office of the Information Commissioner.

The studies cited in support of any provision of pseudoscience were assessed.

### Data Analysis

Descriptive quantitative statistics were derived for: Numbers of NHS trusts identified as offering pseudoscientific therapies; Compliance of the NHS trusts with the FOIA; Compliance of the NHS trusts with standards for evidence based medicine.

Most of the information obtained was only amenable to qualitative processing. The wording of internal NHS policies and processes governing palliative care was extracted and tabulated, where it was related to the scientific basis of the therapy, and its supporting evidence.

The documents received from all trusts as a result of FOIA requests were read in their entirety.

Where trusts provided references to support specific or generic therapies, key data and conclusions were extracted to examine the quality of data. Published papers were found using Google searches, because not all relevant journals are indexed by services such as PubMed. Abstracts provided enough detail to determine key aspects of study design.

Where statements regarding mechanisms and health claims were without reference to published work, implausible quotations were noted where there is no support in the literature. The most implausible mechanisms of action, in relation to established science, were prioritised in this selection.

## Results

### Numbers of Trusts identified as providing pseudoscientific therapies

Of the 25 bodies identified by the initial search, two were not NHS trusts and thus not subject to FOIA. Five trusts no longer offer qualifying therapies, so their websites were out of date. Overall, 19/169 (11%) eligible trusts were found to be offering pseudoscientific practices in oncology and palliative care.

### Responses to FOIA Requests and Compliance with Legislation

All 23 NHS trusts (100%) responded to request for information although this required requests for internal review of the public body’s failure to respond (7/23, 30 %), and complaints to the Office of the Information Commissioner (4/23, 17 %).

The results are shown in Table 1.

**Table 1:** Summary of FOI Requests to NHS Trusts.

| Survey and FOIA request process | N (% , or range ) |
| --- | --- |
| Search hits returned | 25 |
| Body not an NHS trust, not subject to FOIA | 2 |
| Body subject to FOIA, requests submitted | 23 |
| Response received, n (%) | 23 (100) |
| Working days to full response, mean (range) | 29 (2-89) |
| Responses outside statutory limit of 20 days, n (%) | 13 (57) |
| No information on offered therapy held by Trust, n (%) | 6 (17) |
| Therapy no longer provided, n (%) | 5 (22) |

Thirteen NHS trusts received requests that were confined to the provision of reiki. Of these, only three responded within the statutory time limit of 20 working days. Mean response time was 35 working days (range 8-84), the longest being over four months from Cambridge University Hospitals NHS Foundation Trust.

Five trusts stated that no information relevant to the request was held. Of these, one stated that they had never engaged a reiki therapist, so the information on the Sam Buxton charity website was wrong. The remaining four stated that the reason for not holding information was that reiki had been discontinued some years before.

Eight NHS trusts provided at least some correspondence related to the provision of reiki, of which five provided documents governing the practice.

Portsmouth Hospitals University NHS Trust stated that under its record retention policy relevant correspondence had not been retained. University Hospitals Dorset NHS Foundation Trust stated that the cost of retrieving the information would exceed the threshold under the Freedom of Information Act, and was therefore exempt. St Helens and Knowsley Hospitals NHS Trust stated that there were no such records for the last 20 years.

Doncaster and Bassetlaw NHS Foundation Trust stated: “There was no internal guidance located in relation to the provision of the Reiki therapy.” This trust had initially been identified as providing reiki in oncology departments, but on further investigation this was found to be a service for staff not patients.

One trust (Manchester University NHS Foundation Trust) stated: “As outlined in response to Question 1, this post was to be funded fully from external charitable monies. Following this confirmation, the post was approved and advertised through the Trust’s standard recruitment processes.”

Of the governing documents for reiki that were received from five trusts, only two referred to the need for the practice to be evidence based. However no valid evidence was cited. Manchester University NHS Foundation Trust provided a list of references in support of reiki (see table 3, and analysis below).

**Table 2:** Summary of Documents Received: Expanded request.

| Therapy | Count of offers of therapy | Business case provided | Correspondence /meetings | Governance/ guidance | Gov docs require evidence | Evidence for therapy cited or provided<sup>2</sup> | Mode of action justification |
| --- | --- | --- | --- | --- | --- | --- | --- |
| Reiki | 4 | 1 | 0 | 3 | 2 | 1 | 0 |
| Aromatherapy | 6 | 1 | 0 | 5 | 4 | 0 | 0 |
| Emotional freedom technique | 5 | 1 | 0 | 3 | 2 | 0 | 0 |
| Indian head massage | 2 | 1 | 0 | 2 | 1 | 0 | 0 |
| Reflexology | 8 | 0 | 0 | 6 | 4 | 1 | 0 |
| Chakra balancing | 1 | 0 | 0 | 1 | 0 | 0 | 0 |
| Bach flower remedies | 1 | 0 | 0 | 1 | 1 | 0 | 0 |
| Therapeutic touch | 1<sup>3</sup> | 0 | 0 | 0 | 0 | 1 | 0 |
| Total instances | 28<sup>1</sup> | 0 | 0 | 21 | 14 | 3 | 0 |

Based on eight NHS trusts which provided information.
This does not mean that the evidence was robust.
Therapy no longer provided – website out of date.

**Table 3:** Analysis of the Reiki Studies Cited by Manchester University NHS Foundation Trust.

Systematic Reviews
| Setting and/or indication | Conclusion |
| --- | --- |
| Various [11] | “The serious methodological and reporting limitations of limited existing Reiki studies preclude a definitive conclusion on its effectiveness.” |

Randomised Controlled Trials
| Setting and/or indication | Sample size | Blinding | Statistically significant result | Comment |
| --- | --- | --- | --- | --- |
| Fibromyalgia [12] | 93 | Double | No |  |
| Healthy volunteers [13] | 35 | Single | No |  |
| Dementia [14] | 24 | None | Yes |  |
| Acute coronary syndrome [15] | 49 | None | Yes |  |
| Volunteers [16] | 45 | Single | Yes | Small effect size. |
| Cancer pain [17] | 24 | None | Yes | No effect on opioid use. |
| Anxiety, depression, pain [18] | 20 | None | Yes |  |
| Depression and stress [19] | 46 | Patient | Yes |  |

Uncontrolled Studies
| Setting and/or indication | N | Comment |
| --- | --- | --- |
| Occupational health [20] | Not stated | Pilot study on clinicians’ attitudes to reiki |
| Occupational health [21] | 17 |  |
| Physical and psychological trauma [22] | Not stated |  |
| Pain [23] | 20 |  |
| Healthy volunteers [24] | 23 |  |

Non-systematic Reviews and Opinion Pieces
| Setting and/or indication |
| --- |
| Surgery [25] |
| Palliative care [26] |
| Oncology [27] |
| Healing [28] |
| Midwifery [29] |
| HIV/AIDS [30] |

415 Case Reports
| Setting and/or indication |
| --- |
| Pain [31] |
| Dementia [32] |

The correspondence provided by Royal Devon University Healthcare NHS Foundation Trust included a complaint submitted in August 2019 by the National Secular Society, objecting to the provision of reiki. In response, the trust accepted that there was minimal scientific evidence to support the practice, and referred to “a wealth of feedback from patients”.

Data on documents received in response to the expanded request are summarised in table 2. This is based on the information provided in the respective trust’s website. Column 1 shows the frequencies with which therapies were provided. There were 28 instances of the nominated therapies being offered by eight NHS trusts. The leading therapy was reflexology, offered by all eight trusts. For four trusts the websites were out of date and the therapies were no longer provided.

Eight trusts found to be offering pseudoscientific therapies were asked to provide a business case. Only one trust provided a business case, in the form of a funding application for a wellbeing centre at Liverpool University Hospitals NHS Foundation Trust. However, the application was based on perceived patient demand and not on clinical evidence. Four trusts stated that no business case was needed as the service was charity funded, and three trusts could not locate any business case.

Governance documents were provided by five trusts for 21 instances of the therapies offered. Of these, in 14 cases (67%) the document stated that the therapy should be evidence-based. However no trust provided any such evidence for any therapy. Where a mode of action was claimed no justification was provided.

Taking aromatherapy as an example, six trusts offered it, of which none provided a business case or records of correspondence and meetings. Five provided governance documents such as job descriptions or written procedures. Four stated in governance documents that therapies must be evidence based, but none provided or cited such evidence in the documents themselves. Although the trusts made claims about how aromatherapy works, none provided any evidence to justify the claims.

### Evidence for Therapies Provided by Trusts

#### Evidence for specific therapy offered

One NHS trust, Manchester University NHS Foundation Trust, provided a separate list of 22 references to publications which it claimed supported the use of reiki. These are listed in Table 3. The citations comprised: one systematic review; eight randomised controlled trials (RCTs), of which one was double-blind but did not achieve a significant result, three were single-blind, and the remaining four were open-label; and thirteen non-systematic reviews and opinion pieces.

Six of the RCTs yielded a statistically significant result in favour of the test therapy (two single blind, four open label).

No other NHS Trust responded to the request for evidence.

#### Evidence for generic complementary therapy provision

The University Hospitals Plymouth NHS Trust cited six studies, in section 5.2 of their generic Policy for Complementary Therapy Provision as “Examples of Research Evidence”. Table 4 shows the analysis of these six studies. One systematic review and one meta-analysis did not draw positive conclusions. None of the remaining papers cited described a randomised controlled trial.

**Table 4.** Analysis of Relevant Complementary Medicine Studies Cited by University Hospitals Plymouth NHS Trust.

| **Setting and/or therapy** | **Study type** | **N** | **Outcome/Conclusion of the study** |
| --- | --- | --- | --- |
| Spiritual healing [33] | Systematic review | Not stated | Inconclusive |
| General practice [34] | Open label,<br>no comparator | 33 | Qualify of life improved. |
| Spiritual healing [35] | Non-systematic review | Not stated | Calls for systematic research. |
| Therapeutic touch [36] | Meta-analysis | Not stated | Inconclusive |
| Reflexology [37] | Systematic review | Not stated | Probably a non-specific effect. |
| Foot massage in cancer patients [38] | Open label,<br>no comparator | 87 | Further research warranted. |

#### Examples of unreferenced claims

The documents received from all trusts as a result of FOIA requests were read entirely, and statements within them were assessed as to whether they were supported by evidence. Below is listed a non-exhaustive selection of statements for which robust evidence does not exist.

From Barking, Havering and Redbridge University Hospitals NHS Trust:

““Indian head massage relaxes tension in muscle tissue around the head, face, neck, shoulders and arms. By relaxing and stimulating these areas, blood flow increases, and energy balance is restored within the chakras. This helps to reduce stress and anxiety.”

“Flower remedies are now known to help the resolution of stuck emotions and frames of mind. They work on our invisible software, our thoughts and feelings, to retune and rebalance us back to our selves again”.

From The Christie NHS Foundation Trust:

“The treatment combines classic acupressure points with reflex points of the spine, shoulders and legs. In addition, reflexology theory suggests that by working the hands it helps the feet and conversely working the feet may help the hands.”

“Emotional freedom technique is an evidence-based mind-body therapy during which cortisol levels reduce and the brain’s emotional centre becomes less active.”

“Head massage is a treatment that focuses on massaging acupressure points along the head, neck and shoulders…”

“There is no need to remove any clothing as reiki will pass through anything, even plaster casts. The therapist is a channel which the energy is drawn through by the need or imbalance in the individual.”

From University Hospitals Dorset NHS Foundation Trust:

“Chakra Balancing is a therapy which combines aromatherapy, crystal therapy, sound healing and energy healing to work on bringing balance to the chakra system, the seven energy centres located within the body.”

“Our bodies are made up of our physical feelings and sensations and also our energetic frequencies. Chakra Balancing aims to use these different modalities to work on clearing and balancing the energetic frequency surrounding the body and its energy centres.”

From East and North Hertfordshire NHS Trust:

“Energy Therapy is a non invasive therapy in which the therapist uses touch or the placing of hands a short distance from the body, seeking to transfer energy to the recipient in order to bring benefit and a greater sense of well being.”

“Remember – trust the therapy!”

From St Helens and Knowsley Hospitals NHS Trust:

“Through the laying on of hands the therapist will channel energy and unconditional love into the recipient. Reiki healers believe this will balance and energize the mind, body and spirit whilst releasing any energy blockages which may be present enabling the body to be in a receptive state to heal.”

From St Helens and Knowsley Hospitals NHS Trust:

“EFT relieves symptoms by a routine of tapping with the fingertips on a short series of points on the body that correspond to acupuncture points on the energy meridian points stimulating specific parts of the brain, increasing the effectiveness of the energy interventions.”

### Ethics of Therapy Provision

Eleven trusts provided governance documents. All governance documents stated that complementary therapies were not intended to have any effect on the course of disease. They did not state that these therapies had no robust evidence for any other claimed effects.

All specified that patient consent to the therapy proposed was mandatory. Of these, only four specified the information given to patients in relation to obtaining consent. For example, University Hospitals Bristol and Weston NHS Foundation Trust, in its Complementary Therapies Policy, stated that patients were to be given the booklet ‘The Art of Reflexology’. However, this information comes from the Association of Reflexology and hence is not independent.

## Discussion

### Principal Findings

A sizeable proportion of NHS trusts in England offer pseudoscientific therapies in relation to cancer treatment and palliation. Over half of trusts failed to meet their statutory obligation to provide timely responses to FOIA requests. Such information and justification should have been readily available. Almost half of the trusts who responded did not provide any governance documents at all. Only four trusts specified the information on which consent was to be based. It should not be assumed that complementary therapies cannot have any harms, so properly informed consent is a major problem in this field. Although the NHS is under huge pressure to deliver patient care with reduced resources, the FOIA requests were about the governance of patient care and its underpinning evidence. A few governance documents required therapies to be evidence based. However, this was mostly ignored, or the quality of evidence was far from adequate.

No unit claimed any effect of the complementary therapy on the course of disease. Nevertheless, the word ‘healing’ was commonly used, although sometimes qualified by explaining that in this context healing did not mean curing the disease. This is likely to confuse patients, who will understand healing to mean cure, and a different term should be used.

Although one trust did respond to the request for a business case, the funding application provided was not based on research evidence but on what was perceived as patient demand. Citations from two trusts in support of therapies mostly comprised poor quality studies and opinion pieces. This suggests that staff in charge of important decisions do not have a satisfactory grasp of what constitutes evidence, how to read a paper, or evaluate the quality of a study.

### Strengths and Limitations

This innovative survey appears to be the first of its kind, reacting to a story in the national media, and developing iteratively in response to findings. It achieved a high response rate and focussed on a relatively small set of therapies, thus capturing information in depth. Limitations include having no prespecified protocol, exclusion of primary care, and poor knowledge of existent pseudoscientific therapies, so many will have been overlooked thus likely leading to an underestimate of prevalence. Actual patient uptake could not be assessed. Having been triggered by a media story about palliative care, the survey selected NHS units in oncology and related departments. Many search hits were observed for other specialisms. Data were only captured from what trusts had published on their websites. Hence the penetration of pseudoscience into the NHS generally could be far wider.

### Interpretation

Is it really harmful to regale extremely ill people with fictitious stories? The damage immediately relates to misleading patients and abusing their trust. In the future, service providers and other patients may be harmed if therapists who believe that health is governed by a vital force, or by imaginary connections between the feet and other organs, are not challenged, but tolerated by healthcare professionals and encouraged by trusts. Tolerance of pseudoscience is unlikely to encourage adherence to clinical decision-making based on evidence.

There is clearly nothing wrong with giving emotional support to patients with serious physical conditions. However, the publications cited by the NHS trusts in this survey provided no evidence that their therapies ‘work’ in terms of emotional or physical support. The placebo effect is multi- factorial and complex, and can easily explain the positive outcomes which patients report. Patients who experience an improvement in well-being are more likely to be responding to engagement with a caring person who carries out a ritual. It is not necessary to invent mysterious mechanisms in order to achieve these results, which misleads patients, brings dishonesty into the relationship with the healthcare professional, and undermines the principles of evidence-based care.

### Implications for Policy, Practice and Research

There is no convincing evidence for clinical- or cost-effectiveness for these pseudoscientific therapies. Vulnerable patients with cancer are being misled in a sizeable proportion of NHS trusts. Clinicians who work in these services risk having their professionalism compromised. Policy makers should scrutinise more closely what is being offered in the NHS, and specifically avoid the word ‘healing’. The findings should provide further impetus to reassess the evidence base of all clinical practices, to the benefit of patient care and stewardship of the NHS.

### Future Research

This study should be replicated in different settings, countries and time, maybe focusing on different services and other so-called complementary therapies. Qualitative research might explore how these practices became embedded and remain supported. More studies, using different designs should be considered, to assess the evidence base across a range of specialisms.

## Conclusions

Pseudoscientific practices appear to be embedded in England’s National Health Service, and these may undermine progress towards fully evidence-based health care.

## Data Availability

Documents obtained via FOIA requests are available at:

https://www.whatdotheyknow.com/user/les_rose/requests

https://bit.ly/47IrrVs

## Acknowledgment

The author is grateful to Professor Susan Bewley for detailed and constructive criticism of the manuscript.

